# Short-term health system responses to epidemics across hard-to-reach areas in sub-Saharan Africa: A scoping review protocol

**DOI:** 10.1101/2023.05.05.23289564

**Authors:** Annette A. Murunga, Ben Ngoye, Francis P. Wafula, Gilbert O. Kokwaro

## Abstract

**Background:** Epidemics and disease outbreaks are the occurrence of cases of disease in excess of what would be normally expected. Epidemic-prone diseases, including emerging and re-emerging diseases, constitute the greatest threat to public health security and disruption of social and economic development. When cases of epidemics are diagnosed in specific areas, an outbreak response is triggered to stop the spread swiftly, keeping as few people as possible from being infected. In the past 20 years, the sub-Saharan region has witnessed a marked increase in the number of outbreaks in pandemics, such as cholera, dengue, A/H5N 1 influenza, and rift valley fever, among others. And while many of the efforts toward containment have been individually studied, we have no recent studies that examine them collectively in order to draw appropriate comparisons, no recent studies that have especially focused on hard-to-reach areas, and none that have applied a health systems lens. This study thus details a scoping review of short-term health system responses to epidemics across hard-to-reach areas in sub-Saharan Africa.

**Methods:** A scoping review will be undertaken following PRISMA guidelines for scoping reviews. A modified Donabedian framework will be used to help in understanding approaches used in the immediate health system responses to disease outbreaks. The review will focus on published and unpublished studies that report short-term health systems responses during disease outbreaks in sub-Saharan Africa. These will be gleaned from PubMed, google scholar and Cochrane electronic databases, supplemented by a Google advanced search. In addition, manual searches will be carried out through related articles and websites. Two authors will independently screen the studies and extract data. Disagreements will be resolved by discussions, and a third reviewer will decide if there is no consensus. Data will be charted, coded, and narratively synthesized.

**Outcome:** We anticipate developing a document that will show the different approaches health systems responded with to epidemics in the short term and that will contribute to strengthening future short-term responses to disease outbreaks by identifying best practices and innovative ideas. In addition, the study will highlight possible knowledge gaps that need to be filled with new research.

## Introduction

The Covid-19 pandemic pushed the world to the brink of a social and economic meltdown, triggering questions about how well the global health community is prepared to handle major epidemics and pandemics. The world had to deal with an event that has been described as a once-in-a-generation occurrence. That said, we cannot claim full ignorance of the risk of such calamities. Health system inequities and poor resourcing of healthcare, especially across low-and middle-income settings, put the entire world at risk. One type of population group that is particularly ignored and underfunded yet carries a substantial risk of disease outbreaks is those living in hard-to-reach areas across Africa. We define hard-to-reach areas as places where it would be hardest or take the longest for someone to access basic humanitarian services (such as health clinics and hospitals). This access is further compounded by a lack of functioning transport links and infrastructure, as well as terrain difficulty (1).

To contextualize the challenge better, though, it is important to realize that health systems in LMICs are barely able to handle those in high-access areas such as urban and more affluent areas. Major gaps exist in key areas like surveillance, emergency preparedness and response, risk communication and effective control and management of points of entry (2). A WHO evaluation reported that most SSA countries are not equipped to respond to sudden shocks to health systems, yet the region often suffers isolated disease outbreaks (3). Worse still, broader dynamics continue to contribute to the emergence and re-emergence of outbreaks of new diseases and antimicrobial resistance (4). Despite the fact that outbreaks like SARS, cholera, chikungunya, dengue fever, Rift Valley Fever and Ebola provided warning signs and tested countries’ ability to respond (5), Covid-19 still found most LMICs unprepared. And the poor emergency response strategy was made worse by inadequate laboratory and epidemiological surveillance systems and economic, social and political disincentives to case reporting (6).

While Covid-19 presented a particularly unique challenge for most countries, we think that we cannot fully appreciate the path-dependent challenges affecting SSA without going back in time to understand how countries have been responding to other types of outbreaks. We are particularly keen to document immediate (short-term) responses in order to better understand policy agility and capacity to learn, adapt and change when the need does not seem/appear to be urgent. We know that because most SSA countries focus on urgent rather than important matters, their ability to handle Covid-19, which is both an urgent and important matter, was compromised. We argue that by understanding immediate policy response to outbreaks in low-priority areas, we may be able to characterize and deal with the pain points that prevent countries from prioritizing and establishing mechanisms to nip outbreaks in the bud before the major calamity. We also argue that while effort has gone towards examining health sector responses to specific outbreaks, not enough attention has gone towards characterizing commonalities in immediate response to the disease of different kinds and how the immediate actions translate into (or fail to translate into) medium- and longer-term strategies. Examining the causal relationships between the response activities and the broader health system change is crucial if the global health community is to find a lasting solution to disease outbreaks. Moreover, successful strategies for pandemic response and preparedness over the years have been viewed from a vertical lens that is disease-specific, yet it is plausible that if the response was looked at from a health system lens, the response would be quick and more effective. This explicit linkage appears under-addressed by existing literature.

Our scoping review, therefore, aims to provide an overview of existing evidence on immediate health system responses to epidemics in SSA and answer the following questions: what is the context of the health system (Service Delivery, Human Resources, Leadership and Governance, Financing, Medicines Procurement and Supply Chain, and Information Technology) with regard to epidemic preparedness and management in SSA? What is the diversity of approaches in terms of policies, protocols, communication, community participation, and surveillance that were used to manage disease outbreaks? And what were the outcomes of the various approaches and models in the early detection of diseases, reduced disease impact on the population, change in health behaviour, and strengthened health system?

### The rationale for the scoping review

A scoping review is suitable for summarizing and disseminating research findings and identifying research gaps in the existing literature. While a systematic review typically focuses on a well-defined question where appropriate study designs can be identified in advance, a scoping study addresses broader topics where different study designs may be applicable (7).

### Conceptual framework for the scoping review

A conceptual framework has been developed using the Donabedian framework (8) of structure, processes and outcomes (Figure 1). The framework is commonly used to assess the quality of health services. For this review, a modified framework will be used to help in understanding approaches used in the immediate health system responses to disease outbreaks. For the review, ***Structures*** will describe the context of the health system and includes the WHO building blocks of Service Delivery, Human Resource for health (HRH), Leadership Management and Governance (LMG), Financing, Medicines, and Technologies, Health Information Systems (HMIS) (9). ***The process*** will include the functions and the surveillance, policies, protocols, and communication between partners, and ***outcomes*** will include early detection, reduced disease impact on the population and improved health systems.

**Figure 1:**
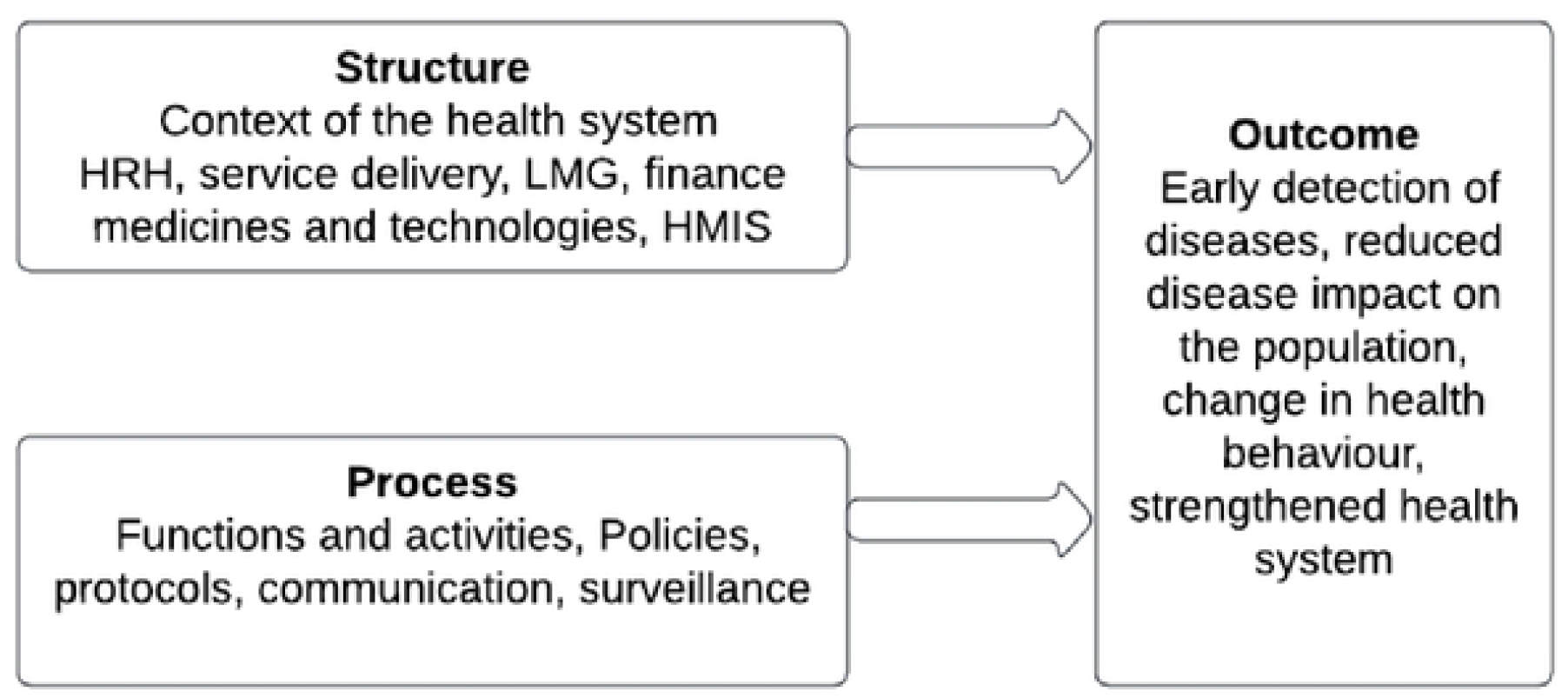
Modified Donabedian framework for the scoping review

**Figure 2.**
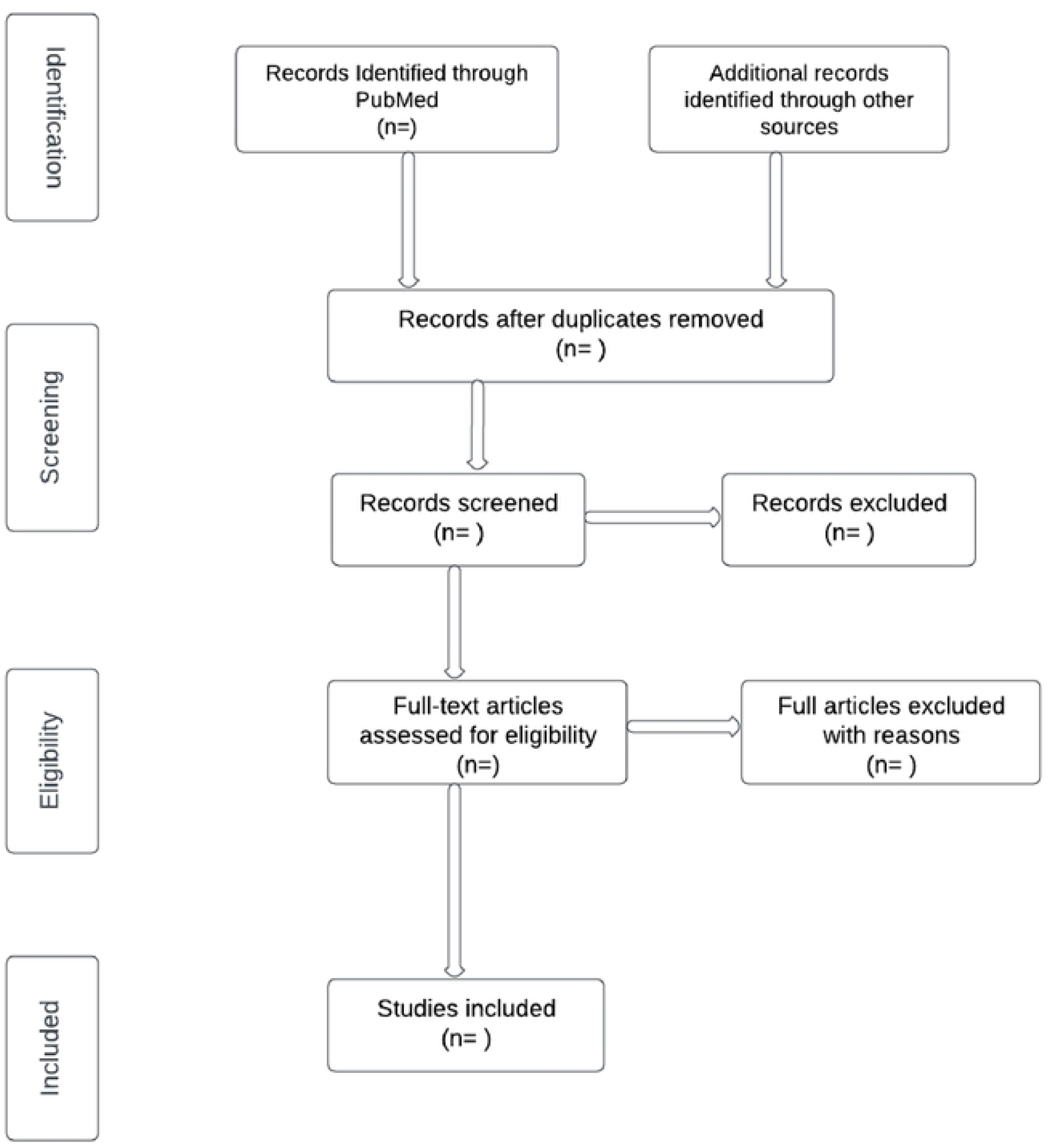
Preferred Reporting Items for Systematic Reviews and Meta-Analyses (PRISMA) flow diagram.

Aided by this modified framework, we focus on the following review questions:

1. Context of the health system building block: *What contextual forces are shaping the health system, and what are the key challenges faced by countries in SSA?*
2. Processes (functions and activities) of the health system: *What are the main functions and activities of the health system in SSA, and what diversity of approaches, models, innovations and tools were used to manage disease outbreaks?*
3. Outcomes of the health system: *How successful are the various approaches and models in improving the prevention and management of disease outbreaks?*

### Methodology

The review seeks to describe the short-term health system responses to epidemics across hard-to-reach areas in sub-Saharan Africa. Like other scoping reviews, the purpose is to broadly describe a phenomenon (what is known and what isn’t) and characterize it in ways that contribute towards a more detailed understanding and/or further research. Consequently, we seek to identify and map available evidence from a wide variety of sources, including published and unpublished material. We follow Arksey and O’Malley’s five-stage scoping review framework (10), which proposes the following five stages: identifying the research question, finding the articles, selecting appropriate articles, charting the data, and collating and summarizing the data into meaningful themes.

#### Stage 1: Identifying the research question

The research question in this review is based on the Joanna Briggs Institute PCC (Population-concept-context) model (11). An overarching research question was developed to guide the search strategy: “What are the short-term health system responses to epidemics across hard-to-reach areas in sub-Saharan Africa?”. The question allows us to capture appropriate existing literature and also provides an opportunity for research questions to be added or modified in the course of the study.

#### Stage 2: Identifying relevant studies

Our approach will include a systematic search of peer-reviewed studies from electronic databases - PubMed, google scholar and Cochrane, snowballing from article reference lists to include additional studies that may not have been indexed. We will also search grey literature from the internet using Google. Literature search strategies will be developed using medical subject headings (MeSH-terms) and text words related to the population, geographical terms and phenomenon of interest/intervention, as shown in Table 1.

**Table 1:**
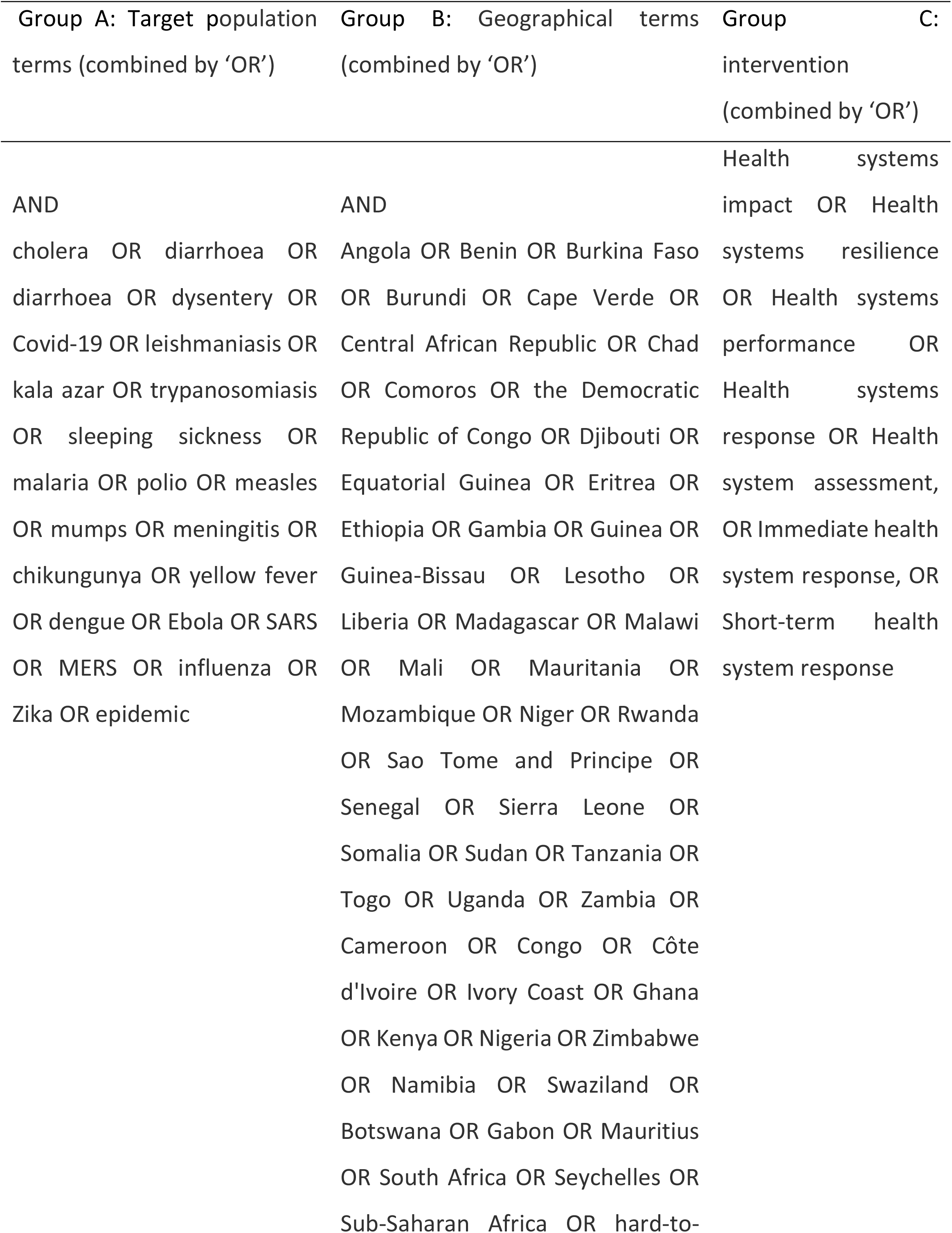
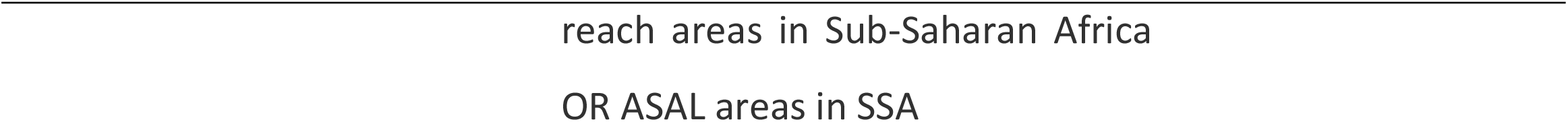
Search terms used in searching Pub-Med electronic database

Empirical studies with either qualitative or quantitative data published in English will be considered for inclusion. The scoping review will exclude all types of reviews, protocols and book chapters. A three-step search strategy will be undertaken. An initial limited search of PubMed, google scholar and Cochrane will be carried out, followed by an analysis of the text words contained in the title and abstract and the index terms used. The second step will include using all identified keywords and index terms in all the included databases. The third step will include screening the reference list of all the studies for additional articles. We shall consider all articles from 2010 onwards.

MeSH terms used to search for CAB Direct included: *Epidemic OR disease outbreak, AND Health system response AND Sub-Saharan Africa*; and for Google Scholar, *Health system response to epidemics AND Sub-Saharan Africa*

#### Stage 3: Selecting articles

Following the search, all identified articles will be uploaded onto Zotero, where all duplicates will be removed. The review process will be carried out in two stages; each stage will have two reviewers who will independently determine the articles based on the inclusion and exclusion criteria. The first screening process will include reading the title and the abstracts to reach the following decisions: (1) if at least one reviewer agrees to include or consider the abstract or title to be inconclusive, the study will be moved to the next level of screening; (2) for any of the studies if both reviewers agree for an article to be excluded, the article will be excluded. All studies at this level will be charted in an Excel sheet.

In the second level of screening, the full texts will be assessed by two independent reviewers. Reasons for any exclusion will be recorded and reported in the scoping review. Any disagreements will be resolved through discussions or with a third reviewer. The results of the search and study selection will be reported in a Preferred Reporting Items for Systematic Reviews and Meta-Analyses (PRISMA) flow diagram.

#### Stage 4: Charting data

To answer the research question, a data charting form in Excel with the following features will be created: authors, year of publication, study title, study design, the aim of the study, country of the study, and key findings relevant to the review objective. A draft charting table is provided (appendix 1). As a preliminary step, the reviewers will independently extract data from the first five articles using the data-charting table and meet to determine whether the approach to data extraction is consistent with the purpose of the study. The draft data extraction tool will be modified and revised as necessary during the extraction process. Modifications will be detailed in the full scoping review article.

#### Stage 5: collating, summarizing and reporting the results

The data from stage 4 will be collated, summarized and reported in a manner that aligns with the study objectives. Tabular and graphical representations of the data may be used to illustrate the identified results and will be supported by narrative descriptions of the data.

The meaning of the findings and implications for future research and practice will be discussed.

#### Patient and public involvement

Patients and the public will not be involved in the design and conception of the study.

#### Ethics and dissemination

The proposed scoping review does not require ethical approval as data will be collected through published peer-reviewed literature and grey literature. This scoping review will provide a comprehensive overview of existing evidence in the field.

## Data Availability

No datasets were generated or analysed during the current study. All relevant data from this study will be made available upon study completion.

---

**Appendix 1:**
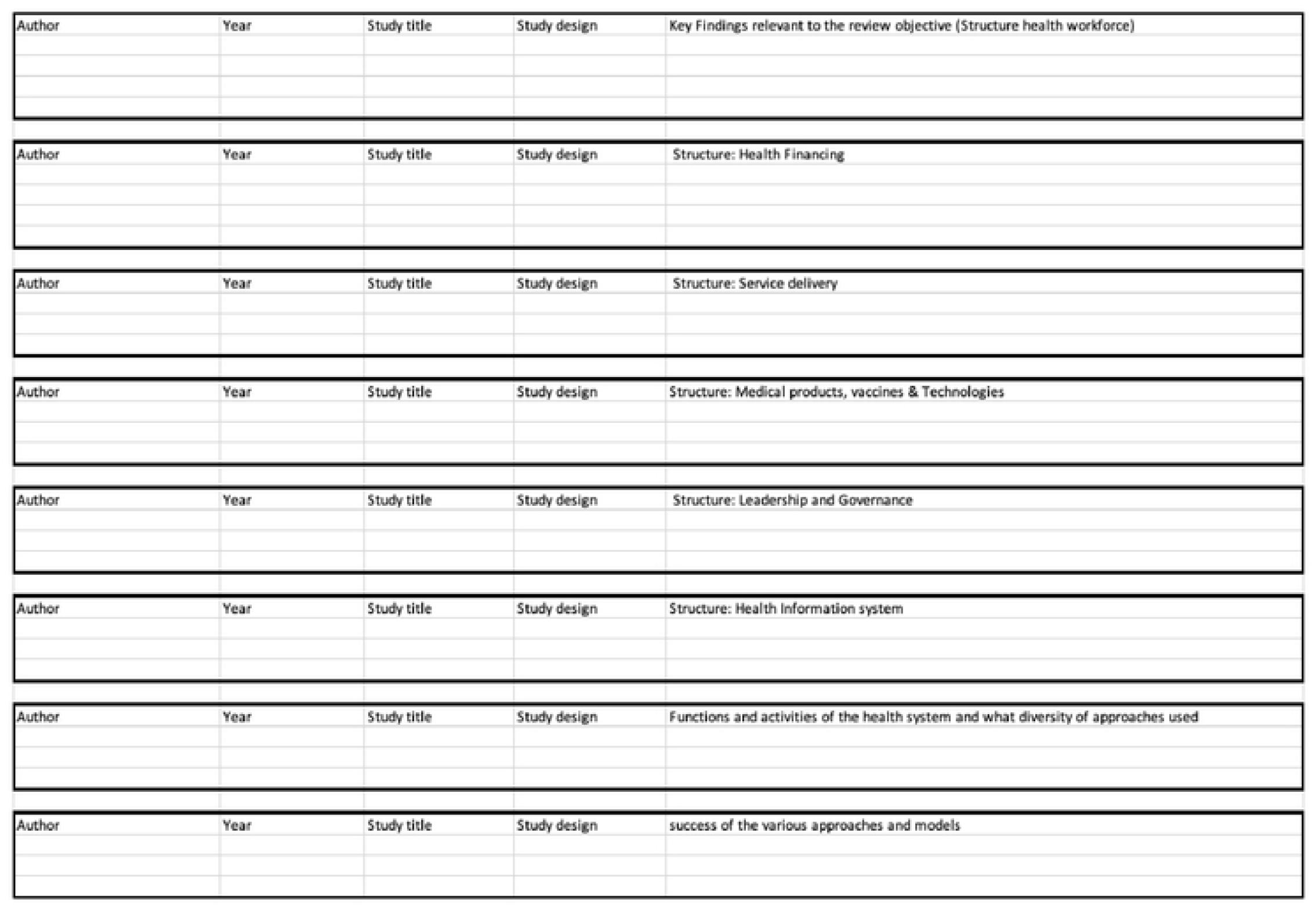
Data extraction sheet

